# Evaluation of the SEIMC score as a death predictor of SARS-CoV-2 pneumonia in vaccinated patients: a validation cohort

**DOI:** 10.1101/2025.07.09.25331178

**Authors:** Jaime Gil-Rodríguez, Pablo Aranda-Laserna, Miguel Ángel Montero-Alonso, Javier Martínez de Victoria Carazo, Daniel Fernández Reyes, Emilio Guirao-Arrabal

**Affiliations:** Internal Medicine Unit. San Cecilio University Hospital. Avenida del Conocimiento s/n, 18016, Granada. Spain; Department of Statistics and Operations Research, University of Granada. Granada. Spain; Infectious Diseases Unit. San Cecilio University Hospital. Avenida del Conocimiento s/n, 18016, Granada. Spain

**Keywords:** SARS-CoV-2, COVID-19, COVID-19 vaccines

## Abstract

**Introduction:** This is a single-centre prospective cohort study to validate the Spanish Society of Infectious Diseases and Microbiology (SEIMC) score for predicting 28-days mortality in vaccinated patients with SARS-CoV-2 pneumonia.

**Methods:** We included 127 adult patients with confirmed SARS-CoV-2 infection by polymerase chain reaction (PCR) admitted to the hospital ward. Among them, 92 were unvaccinated, 17 one-dose vaccinated and 18 two-doses. The SEIMC score was calculated at admission, and we analysed its correlation with 28-days mortality.

**Results:** We found significant differences between the number of vaccinations according to the SEIMC score classification. The SEIMC score and 28-days mortality were significantly correlated in the whole cohort (p=0.001), in unvaccinated patients (p=0.001) and in those with some vaccine doses (p=0.003) or two-doses (p = 0.005), but not in one-dose vaccinated patients (p=0.235).

**Conclusion:** The SEIMC score remains a useful scale for predicting 28-day mortality in vaccinated patients.

## Introduction

The major milestone of the SARS-CoV-2 pandemic was the initiation of mass vaccination. From the first human vaccine trial in March 2020 to February 2022, 26 vaccines have been approved, with 121 candidates currently in clinical trials and 710 million doses having been administered from December 2020 to 7 April 2021^1,2^. Despite the variability of vaccine effectiveness depending on multiple factors, this has changed the epidemiology of the disease, reducing mortality and the need for intensive care^3,4^.

However, one of the challenges of the pandemic is to predict which patients would develop severe disease given the wide clinical variability of SARS-CoV-2 infection^5^. To this end, multiple risk prediction models have been designed. One of the most relevant scores developed in Europe is the designed by the Spanish Society of Infectious Diseases and Clinical Microbiology (SEIMC score). This score is based on two large retrospective cohorts of hospitalised COVID-19 patients in Spain in 2020 between February and April 2020, months before mass vaccination was approved in Spain^6^.

This raises the question of whether these tools developed in a non-vaccine environment are still valid in the present situation. Currently, 80.9% of the Spanish population has received the full vaccination schedule by 10 February 2022^7^. Hence, we performed a study investigating the utility of the SEIMC score to predict 28-days mortality in vaccinated patients with SARS-CoV-2 pneumonia.

## Methods

We conducted a single-centre prospective cohort study at Hospital Universitario Clínico San Cecilio (Granada, Spain), developed for the study of different prognostic scores such as the Lung Ultrasound Score (LUS)^8^. The study’s protocol was approved by the regional ethics committee with the code 0259-N-21, and all patients included in the study have given their written informed consent. This study followed the TRIPOD guidelines for risk prediction models^9^.

This study included adults with confirmed SARS-CoV-2 infection by polymerase chain reaction (PCR) within the first 24 hours of admission from 01 January 2021 to 14 September 2021^10^. All patients with suspected bacterial superinfection and those who were unable to cooperate or who voluntarily refused to be included in the study were excluded.

The Emergency Department requested analytical data at first contact to the hospital, and the missing data were collected at the time of the examination by the researchers on the COVID-19 transitional hospitalisation ward. This information was used to calculate the SEIMC score within the first 24 hours after hospitalisation, according to the following items: age, age-adjusted O_2_ saturation (SpO2, equal or less than 90% for patients older than 50 years and equal or less to 93% in patients younger or equal to 50 years), neutrophil-to-lymphocyte ratio (NLR), estimated glomerular filtration rate (eGFR) by the CKD-EPI formula^11^, presence or absence of dyspnea and patient’s sex. These variables were used to determine the score, in a range from 0 to 30. Based on it, patients were classified into four risk groups: low risk (from 0 to 2 points), moderate risk (from 3 to 5 points), high risk (from 6 to 8 points) and very high risk (from 9 to 30 points). The outcome measured was 28-days all-cause mortality, and patients that were discharged alive before 28 days after admission were assumed to have survived for at least 28 days.

According to Shapiro Wilks test, the sample was not normal. Therefore, we used Chi-square tests to analyse the score based on 28-days mortality and the differences between the number of vaccinations and severity classification in SEIMC score. To analyse differences based on the number of vaccinations, we used the ANOVA test. Finally, the Bonferroni test was used to analyse differences among subgroups.

## Results

1.205 patients were admitted to the COVID-19 ward of the San Cecilio University Hospital between 1st January and 12th August 2021 with positive SARS-CoV-2 PCR. 1102 of them presented COVID-19 pneumonia and 310 met the exclusion criteria. Finally, 127 consecutive patients were included in the study out of the 718 eligible candidates, based on the availability of human and material resources. Between them, 92 were unvaccinated, 17 had received one dose of vaccination, and 18 had received two doses. The patient selection process is described in Figure 1.

**Figure 1.**
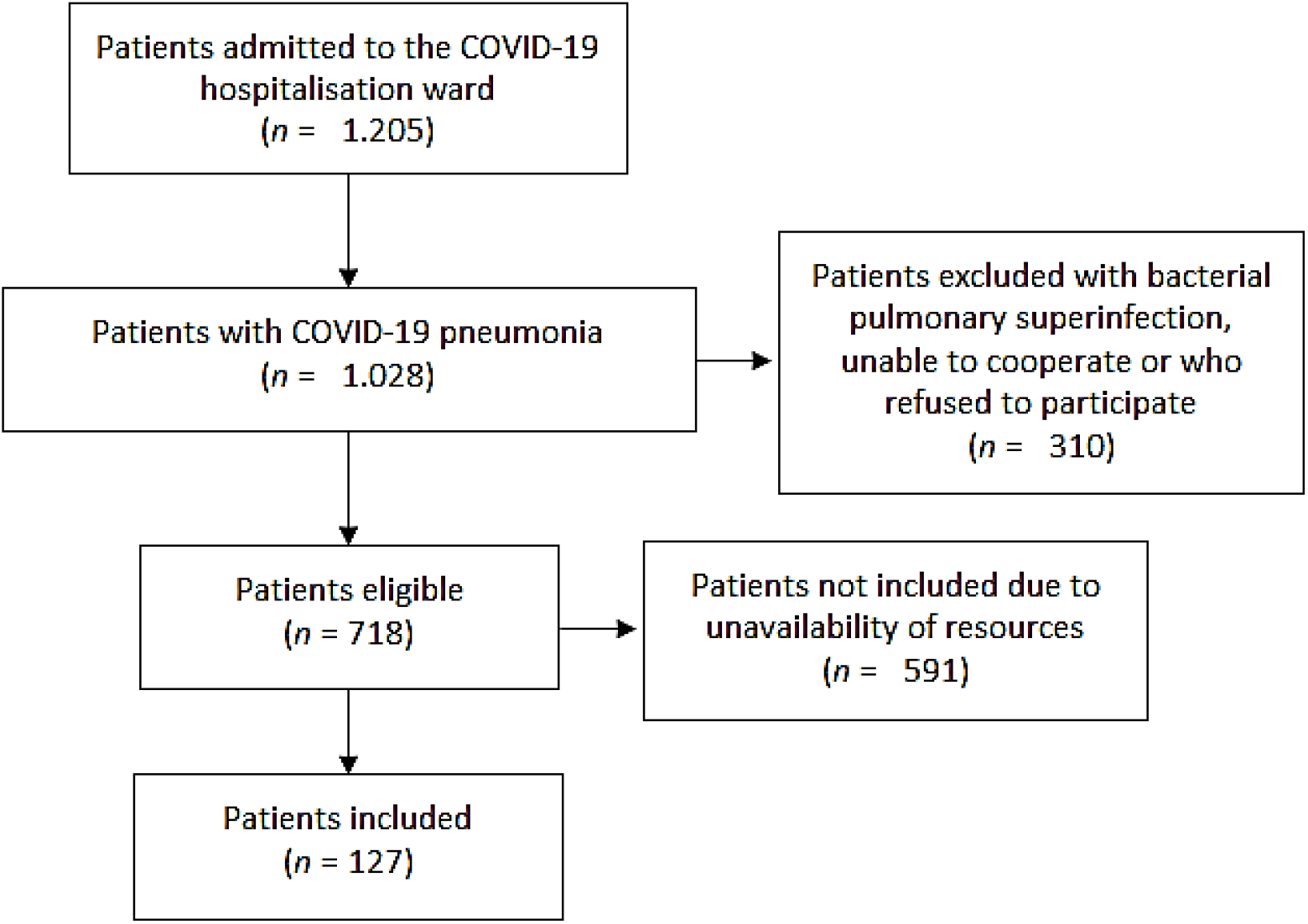
Patient selection flow diagram

The baseline characteristics of the patients are described in Table 1. Median (m) age was 61 years (interquartile range [IQR] 48-73), being significantly higher (p = 0.001) in the two-doses subgroup (m 75, IQR 40-89) than in unvaccinated (m 59, IQR 48-71) or one-dose (m 60, IQR 21-90); and the majority of participants were males (63%), with no statistically significant differences between them. The two-doses subgroup also showed a higher number of comorbidities, and a statistically significant greater proportion of patients with chronic obstructive pulmonary disease, chronic renal disease and hypertension. No differences were found between subgroups in the incidence of dyspnoea, SpO2, need for supplemental O2 or PaO2-FiO2 ratio. Nor in the analytical values of aspartate aminotransferase (AST), lactate dehydrogenase (LDH) or C-reactive protein (CRP). However, there were statistically significant differences in eGFR and NLR between the two-dose vaccine group (m and SD 55.4, 30.0 and 10.5, 10.5 respectively), compared to the unvaccinated (81.4, 25.3 and 6.1, 4.1) and one-dose vaccine (83.6, 21.5 and 6.0, 2.9) subgroups.

**Table 1.** Patients baseline characteristics.

|  | All<br>patients<br>(n = 127) | 0<br>vaccines<br>(n = 92) | 1<br>vaccine<br>(n = 17) | 2<br>vaccines<br>(n = 18) | p-<br>value |
| --- | --- | --- | --- | --- | --- |
| **Age** | 61 | 59.5 | 60 | 75 | 0.001 |
| (median, IQR) | (48-73) | (48-71) | (21-90) | (40-89) |  |
| **Sex** |  |  |  |  |  |
| Male (%) | 80 (63.0) | 60 (47.2) | 11 (8.7) | 9 (7.1) | 0.468 |
| Female (%) | 47 (37.0) | 32 (25.2) | 6 (4.7) | 9 (7.1) |  |
| **Weight [kg]** | 85 | 85.2 | 85.1 | 83.6 | 0.955 |
| (mean, SD) | (18.7) | (18.0) | (14.0) | (26.9) |  |
| **Height [cm]** | 168.2 | 169.1 | 171.6 | 158.7 | 0.009 |
| (mean, SD) | (13.1) | (10.4) | (9.7) | (24.4) |  |
| **Dementia (%)** | 4 (3.1) | 3 (2.4) | 0 | 1 (0.8) | 0.638 |
| **COPD<sup>1</sup> (%)** | 10 (7.9) | 5 (3.9) | 0 | 5 (3.9) | 0.002 |
| **Chronic renal disease (%)** | 18 (14.2) | 11 (8.7) | 1 (0.8) | 6 (4.7) | 0.034 |
| **Diabetes mellitus (%)** | 31 (24.4) | 22 (17.3) | 3 (2.4) | 6 (4.7) | 0.546 |
| **Hypertension (%)** | 59 (46.5) | 40 (31.5) | 5 (3.9) | 14 (11.0) | 0.009 |
| **Obesity (%)** | 46 (36.2) | 33 (26.0) | 5 (3.9) | 8 (6.3) | 0.862 |
| **Neoplasia (%)** | 15 (11.8) | 12 (9.4) | 1 (0.8) | 2 (1.6) | 0.699 |
| **Cerebrovascular disease (%)** | 10 (7.9) | 5 (3.9) | 1 (0.8) | 4 (3.1) | 0.051 |
| **Cardiovascular disease (%)** | 8 (6.3) | 7 (5.5) | 0 | 1 (0.8) | 0.490 |
| **Heart failure (%)** | 7 (5.5) | 7 (5.5) | 0 | 0 | 0.244 |
| **Immunosuppression (%)** | 5 (3.9) | 5 (3.9) | 0 | 0 | 0.372 |
| **Number of comorbidities** | 1 | 1 | 0 | 3 | <0.001 |
| (median, IQR) | (0-5) | (0-4) | (0-5) | (0-5) |  |
| **Days of symptoms** | 7 | 8 | 6.5 | 6.5 | 0.957 |
| (median, IQR) | (2-21) | (2-18) | (3-15) | (2-21) |  |
| **Dyspnea (%)** | 88 (69.3) | 67 (52.8) | 10 (7.9) | 11 (8.7) | 0.371 |
| **Cough (%)** | 90 (70.9) | 68 (53.5) | 12 (9.4) | 10 (7.9) | 0.293 |
| **Altered consciousness (%)** | 4 (3.1) | 3 (2.4) | 0 | 1 (0.8) | 0.638 |
| **Fever (%)** | 48 (37.8) | 35 (27.6) | 10 (7.9) | 3. (2.4) | 0.037 |
| **SaO<sub>2</sub><sup>2</sup> [%]** | 96.2 | 96.3 | 96.4 | 95.8 | 0.731 |
| (mean, SD) | (2.2) | (2.3) | (1.7) | (2.3) |  |
| **Supplementary O<sub>2</sub> (%)** | 76 (59.8) | 57 (44.9) | 10 (7.9) | 9 (7.1) | 0.636 |
| **RR<sup>3</sup> [breaths/min]** | 22 | 23 | 24 | 20 | 0.424 |
| (median, IQR) | (12-38) | (12-38) | (14-28) | (14-32) |  |
| **HR<sup>4</sup> [beats/min]** | 81 | 81 | 82 | 81.5 | 0.784 |
| (median, IQR) | (50-133) | (53-115) | (56-133) | (50-100) |  |
| **SBP<sup>5</sup> (mmHg)** | 127.3 | 126.6 | 126.3 | 132.3 | 0.338 |
| (mean, SD) | (16.4) | (15.0) | (18.1) | (21.0) |  |
| **CT PTE<sup>7</sup> (%)** | 8 (6.3) | 7 (5.5) | 0 | 1 (0.8) | 0.490 |
| **PaO<sub>2</sub> – FiO<sub>2</sub><sup>8</sup> ratio** | 371.5 | 371.1 | 392.8 | 353.8 | 0.561 |
| (mean, SD) | (107.1) | (105.2) | (92.9) | (130.0) |  |
| **Glucose [mg/dL]** | 139.4 | 142.9 | 128.8 | 131.6 | 0.557 |
| (mean, SD) | (59.0) | (64.4) | (33.2) | (48.3) |  |
| **eGFR<sup>9</sup>** | 78.0 | 81.4 | 83.6 | 55.4 | <0.001 |
| [ml/min/1.73m<sup>2</sup>]<sup>9</sup> | (27.0) | (25.3) | (21.5) | (30.0) |  |
| **AST<sup>10</sup> [U/L]** | 50.2 | 53.8 | 44.2 | 37.3 | 0.352 |
| (mean, SD) | (47.6) | (54.1) | (22.1) | (19.9) |  |
| **Ferritin**[ng/mL] | 703.4 | 802.7 | 527.7 | 383.7 | 0.012 |
| (mean, SD) | (610.9) | (679.7) | (308.2) | (187.2) |  |
| > 700 ng/mL (%) | 44 (34.6) | 40 (31.5) | 4 (3.1) | 0 | 0.001 |
| **LDH**<sup>11</sup> [U/L] | 333.2 | 335.5 | 353.1 | 302.2 | 0.397 |
| (mean, SD) | (114.8) | (118.1) | (107.0) | (104.0) |  |
| > 400 U/L (%) | 27 (21.3) | 21 (16.5) | 5 (3.9) | 1 (0.8) | 0.177 |
| **CRP**<sup>12</sup> [mg/L] | 93.4 | 90.6 | 95.8 | 105.4 | 0.629 |
| (mean, SD) | (60.0) | (64.3) | (39.7) | (53.3) |  |
| > 15 mg/mL (%) | 120 (94.5) | 86 (67.7) | 16 (12.6) | 18 (14.2) | 0.539 |
| **Lymphocyte** [10<sup>3</sup>/μL] | 0.9 | 0.9 | 1.0 | 1.2 |  |
| (mean, SD) | (0.6) | (0.4) | (0.4) | (1.1) | 0.095 |
| **Neutrophil - lymphocyte ratio** | 6.7 | 6.1 | 6.0 | 10.5 | 0.007 |
| (mean, SD) | (5.5) | (4.1) | (2.9) | (10.5) |  |
| > 10 (%) | 21 (16.5) | 14 (11.0) | 2 (1.6) | 5 (3.9) | 0.360 |
| **Platelets** [10<sup>3</sup>/μL] | 190.3 | 197.1 | 171.7 | 173.2 |  |
| (mean, SD) | (70.3) | (72.2) | (67.6) | (59.6) | 0.212 |
| **D dimer** [mg/L] | 3.5 | 4.3 | 0.7 | 2.3 | 0.804 |
| (mean, SD) | (22.0) | (26.0) | (0.3) | (3.0) |  |
| > 1,5 mg/mL (%) | 25 (19.7) | 20 (15.7) | 0 | 5 (3.9) | 0.076 |
| **ECO LUS total** | 15.3 | 15.2 | 15.9 | 14.9 | 0.841 |
| (median, IQR) | (5.5) | (5.8) | (4.5) | (5.2) |  |
| > 15 (%) | 65 (51.2) | 49 (38.6) | 9 (7.1) | 7 (5.5) | 0.530 |

<sup>1</sup>**COPD**: Chronic obstructive pulmonary disease; <sup>2</sup>**SaO<sub>2</sub>**: Peripheral arterial oxygen saturation; <sup>3</sup>**RR**: Respiration rate; <sup>4</sup>**HR**: Heart rate; <sup>5</sup>**SBP**: Systolic blood pressure; <sup>6</sup>**RALE**: Radiographic Assessment of Lung (O)Edema; <sup>7</sup>**CT PTE**: Computer Tomography Pulmonary Thromboembolism; <sup>8</sup>**PaO<sub>2</sub> – FiO<sub>2</sub>**: Arterial oxygen pressure to oxygen inspired fraction ratio; <sup>9</sup>**eGFR**: Estimated glomerular filtration rate calculated by the CKD-EPI; <sup>10</sup>**AST**: Aspartate aminotransferase; <sup>11</sup>**LDH**: Lactate dehydrogenase; <sup>12</sup>**CRP**: C-reactive protein; <sup>13</sup>**NT-proBNP**: N-terminal pro-brain natriuretic peptide.

The mean SEIMC score was 7.2 in unvaccinated (SD 4.84, 95% confidence interval [95% CI] 6.19 - 8.20), 6.88 in one-dose vaccine (SD 5.797, 95% CI 3.90 - 9.86) and 12.28 in two-doses vaccine (SD 5.582, 95% CI 9.50 - 15.05). 10.2% of patients showed low risk (11.96% unvaccinated, 11.76% in one-dose, 0% in two-doses), 31.5% moderate (34.78%, 35.29% and 11.11%), 25.2% high (25%, 35.29% and 16.67%) and 33.1% very high (28.26%, 17.65% and 72.22%); with significant differences between number of vaccinations according to risk classification (p = 0.012). These results are explained in Table 2.

**Table 2.**
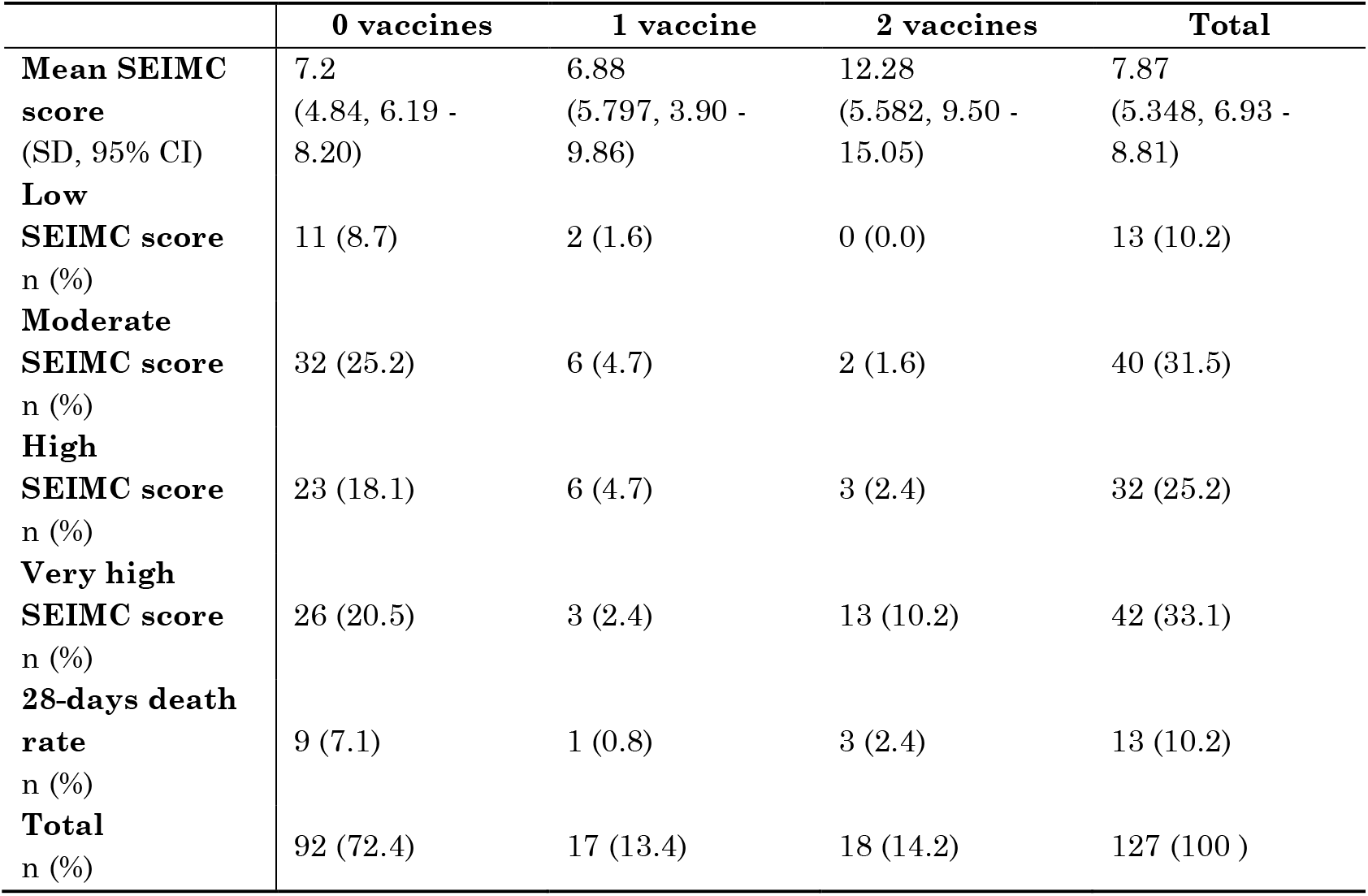
SEIMC score results by number of vaccinations and risk classification.

According to the analysis, the SEIMC score and 28-days mortality were significantly correlated in the whole cohort (p = 0.001), in unvaccinated patients (p = 0.001), in those with some vaccine doses (p = 0.003), and two-doses (p = 0.005), but not in those with one-dose (p = 0.235). Based on the SEIMC score, significant differences were observed between the unvaccinated and those with two vaccinations (p = 0.001), as well as between the one-dose group and the two-dose group (p = 0.006).

## Discussion

This is the first study that has compared the SEIMC score in vaccinated and unvaccinated patients. This score has been externally validated in the general population, in emergency department patients and in more specific populations, such as kidney transplant recipients^12–14^. However, its performance in vaccinated patients has not been evaluated in other studies yet.

The present study was conducted in a dynamic cohort, which has been evaluating patients in the successive waves of the pandemic. This fact may explain the higher number of severe patients in the unvaccinated patients (when hospitals were overwhelmed and only the most severe patients were admitted) and those with two-doses (which corresponds to the outbreaks at the end of summer 2021) than in patients with one-dose of vaccine (where the nadir of admissions occurred in 2021, and less severe patients could be treated). Our results showed significant differences between the number of vaccinations according to the SEIMC score risk classification, which supports this hypothesis. These differences are especially remarkable for the variables age, eGFR and NLR, which justify the differences between risk groups.

The analysis of the correlation between the SEIMC score and 28-days mortality confirmed the good performance of this score in our cohort. This correlation is significant both in unvaccinated patients and, for the first time, in patients with some dose of vaccine. In the subgroup analysis, however, a significant correlation is observed in patients with two-doses of vaccine, but not in those with one-dose. This may correspond, as previously described, to the lower number of severe patients included in this subgroup of patients.

The results of this study provide additional evidence to the published literature on the use of prognostic scores in COVID-19 in vaccinated patients. The main limitation of this study is the sample size, which has made it difficult to establish a statistically significant correlation for some data that showed a positive trend (such as the subgroup of those vaccinated with one-dose).

## Conclusion

The SEIMC score remains a useful scale for predicting 28-day mortality in vaccinated patients.

## Data Availability

All data produced in the present work are contained in the manuscript

## Acknowledgements

To Valme Sánchez Cabrera for its management of the database, and to Alejandra Ciria García for the language supervision of the text.

## References

1. The London School of Hygiene & Tropical Medicine Vaccine Center . COVID-19 vaccine tracker. https://vac-lshtm.shinyapps.io/ncov_vaccine_landscape/. Accessed February 18, 2022.

2. Mathieu E, Ritchie H, Ortiz-Ospina E, et al. A global database of COVID-19 vaccinations. Nat Hum Behav. 2021;5(7):947–953. doi:10.1038/s41562-021-01122-8

3. Tregoning JS, Flight KE, Higham SL, Wang Z, Pierce BF. Progress of the COVID-19 vaccine effort: viruses, vaccines and variants versus efficacy, effectiveness and escape. Nat Rev Immunol 2021 2110. 2021;21(10):626–636. doi:10.1038/s41577-021-00592-1

4. Haas EJ, Angulo FJ, McLaughlin JM, et al. Impact and effectiveness of mRNA BNT162b2 vaccine against SARS-CoV-2 infections and COVID-19 cases, hospitalisations, and deaths following a nationwide vaccination campaign in Israel: an observational study using national surveillance data. Lancet. 2021;397(10287):1819–1829. doi:10.1016/S0140-6736(21)00947-8

5. Dorjee K, Kim H, Bonomo E, Dolma R. Prevalence and predictors of death and severe disease in patients hospitalized due to COVID-19: A comprehensive systematic review and meta-analysis of 77 studies and 38,000 patients. PLoS One. 2020;15(12):e0243191. doi:10.1371/JOURNAL.PONE.0243191

6. Berenguer J, Borobia AM, Ryan P, et al. Development and validation of a prediction model for 30-day mortality in hospitalised patients with COVID-19: The COVID-19 SEIMC score. Thorax. 2021;0:1–10. doi:10.1136/thoraxjnl-2020-216001

7. Ministerio de Sanidad. Cuadro de mando resumen de datos de vacunación. https://www.sanidad.gob.es/profesionales/saludPublica/ccayes/alertasActual/nCov/pbiVacunacion.htm. Accessed February 18, 2022.

8. Gil-Rodríguez J, Martos-Ruiz M, Peregrina-Rivas J-A, et al. Lung Ultrasound, Clinical and Analytic Scoring Systems as Prognostic Tools in SARS-CoV-2 Pneumonia: A Validating Cohort. Diagnostics. 2021;11(12):2211. doi:10.3390/diagnostics11122211

9. Collins GS, Reitsma JB, Altman DG, Moons KGM. Transparent Reporting of a multivariable prediction model for Individual Prognosis Or Diagnosis (TRIPOD): The TRIPOD Statement. Br J Surg. 2015;102(3):148–158. doi:10.1002/bjs.9736

10. Kevadiya BD, Machhi J, Herskovitz J, et al. Diagnostics for SARS-CoV-2 infections. Nat Mater. 2021;20(5):593–605. doi:10.1038/s41563-020-00906-z

11. Levey AS, Stevens LA, Schmid CH, et al. A New Equation to Estimate Glomerular Filtration Rate. Ann Intern Med. 2009;150(9):604. doi:10.7326/0003-4819-150-9-200905050-00006

12. Lombardi Y, Azoyan L, Szychowiak P, et al. External validation of prognostic scores for COVID-19: a multicenter cohort study of patients hospitalized in Greater Paris University Hospitals. Intensive Care Med. 2021;47(12):1426–1439. doi:10.1007/S00134-021-06524-W/TABLES/5

13. Hohl CM, Rosychuk RJ, Archambault PM, et al. The CCEDRRN COVID-19 Mortality Score to predict death among nonpalliative patients with COVID-19 presenting to emergency departments: a derivation and validation study. Can Med Assoc Open Access J. 2022;10(1):E90–E99. doi:10.9778/CMAJO.20210243

14. Modelli de Andrade LG, de Sandes-Freitas TV, Requião-Moura LR, et al. Development and validation of a simple web-based tool for early prediction of COVID-19-associated death in kidney transplant recipients. Am J Transplant. 2022;22(2):610–625. doi:10.1111/AJT.16807

